# SARS-CoV-2 infection drives a glycan switch of peripheral T cells at diagnosis

**DOI:** 10.1101/2021.02.17.21251918

**Authors:** Inês Alves, Manuel M. Vicente, Joana Gaifem, Ângela Fernandes, Ana M. Dias, Cláudia S. Rodrigues, José Carlos Oliveira, Nair Seixas, Luis Malheiro, Miguel A. Abreu, Rui Sarmento e Castro, Salomé S. Pinho

## Abstract

COVID-19 is a highly selective disease in which SARS-CoV-2 infection can result in different clinical manifestations ranging from asymptomatic/mild to severe disease that requires hospitalization. Here, we demonstrated that SARS-CoV-2 infection results in a glycosylation reprogramming of circulating lymphocytes at diagnosis. We identified a specific glycosignature of T cells, defined upon SARS-CoV-2 infection and apparently triggered by a serological factor. This specific glycan switch of T cells is detected at diagnosis being more pronounced in asymptomatic patients. We further demonstrated that asymptomatic patients display an increased expression of a viral-sensing receptor, through the up-regulation of DC-SIGN in monocytes. We showed that higher levels of DC-SIGN in monocytes at diagnosis correlates with better COVID-19 prognosis. These new evidences pave the way to the identification of a novel glycan-based response in T cells that may confer protection against SARS-CoV-2 infection in asymptomatic patients, highlighting a novel prognostic biomarker and potential therapeutic target.

## Introduction

Coronavirus SARS-CoV-2 is the etiologic agent responsible for the global pandemic of Coronavirus disease 2019 (COVID-19), that, as of 9^th^ of February, has infected over 106 million people worldwide(1, 2). The overall mortality of COVID-19 is between 0.5% and 3.5%(1, 2). COVID-19 is a highly selective disease. In fact, only some infected individuals get sick, and although most of the critically ill are elderly, some patients that die are previously healthy and/or relatively young(3). There is an urgent need to improve the understanding of the pathophysiology of this disease, envisioning better management in terms of patients care, treatment options, vaccination strategies and the allocation of healthcare resources. Vaccine development against SARS-CoV-2 poses a great promise for the resolution of the pandemic, and several vaccines are now being distributed among the world population, namely mRNA- and protein-based approaches(4, 5). However, its long-term protection, effectiveness to re-infection, efficacy to new variants and availability for the general public is still on trial. This lack of knowledge highlights the importance of understanding patient-specific immune response to infection, envisioning the identification of novel mechanisms of disease. Moreover, comprehensive insights on COVID-19 molecular mechanism may be translated into diagnostic or prognostic biomarkers, therapeutic targets to improve patient’s clinical management and patients’ stratification for vaccination.

SARS-CoV-2 infection results in a broad range of symptoms(6). Several studies on immune profiling of infected patients consistently revealed an immunological dysregulation, observed in peripheral blood mononuclear cells (PBMCs). A decrease of T cells and dendritic cells frequencies and an increase of monocytes and neutrophils have been observed in hospitalized infected individuals, as well as a cytokine dysregulation, the so-called “cytokine storm” associated with critical illness(7–10). However the mechanism underlying this immunological dysregulation remains largely unknown.

Despite overlooked, the field of glycobiology can provide missing answers to immunological questions. Glycosylation is a major post-translational mechanism characterized by the enzymatic addition of glycans (carbohydrates) to proteins or lipids of essentially all cells, including immune cells. In fact, glycans are master regulators of immune cell functions, defining the activation and differentiation of T cells(11, 12). In the cellular immune system, we and others have previously demonstrated the regulatory power of glycans in adaptive immune response through the modulation of T cell activation thresholds associated with the immunopathogenesis of autoimmune diseases and cancer(13–19). Moreover, being present in several pathogens, glycans can also be sensed by immune cells, through their recognition by specific glycan-recognizing receptors expressed in those cells(20). Like the majority of viruses, SARS-CoV-2 viral capsids are also covered with glycans. More specifically, the spike protein of the SARS-CoV-2 envelope was shown to be highly glycosylated (harboring 22 *N*-glycosylation sites) with glycan structures such as oligomannose and some branched *N*-glycans(21, 22). These structures can be specifically recognized by glycan binding proteins (GBPs) of host’s immune cells, such as the C-type-lectin Dendritic Cell-Specific Intercellular adhesion molecule-3-Grabbing Non-integrin (DC-SIGN), present in innate immune cells, promoting virus recognition and elimination(23, 24).

Defining the glycosylation profiles of immune cells in SARS-CoV-2 infected individuals and how they impact their effector functions remains completely unknown, being of utmost importance for the understanding of COVID-19 immunopathogenesis, as well as for the improvement of the clinical management of the disease and for pandemic control measures, namely as a stratification biomarker.

In this study, we discovered that circulating T cells exhibit a glycan switch upon SARS-CoV-2 infection that is detected at COVID-19 diagnosis. This change in the glycosylation profile of T cells appears to be triggered by a serum inflammatory factor present in infected patients. This specific T cell glycan switch is more pronounced in asymptomatic patients, rather than in those who exhibit symptoms. Importantly from a clinical standpoint, we also unveiled that COVID-19 patients with good prognosis exhibit an upregulation of DC-SIGN expression in circulating monocytes.

## Material and Methods

### Cohort description and patient’s selection criteria

The present study integrates a total of 32 patients diagnosed with SARS-CoV-2 from 2 individual Portuguese cohorts (between May 2020 and July 2020), 20 patients from Infectious Disease Department, Centro Hospitalar e Universitário do Porto (CHUP), Porto, Portugal and 12 patients from Infectious Disease Department, Centro Hospitalar de Vila Nova de Gaia/Espinho (CHVNG), Vila Nova de Gaia, Portugal. The total cohort includes patients of both genders and age between 22 and 89 years old. All the samples were included in all the FACS analysis discussed.

Blood was collected and plasma and PBMCs were isolated at the time of diagnosis and 14 days (n=6) after diagnosis. Similar analysis was conducted in a subset of patients (n=2) that recovered.

The eligibility criteria for inclusion in this study were SARS-CoV-2 positive patients asymptomatic or symptomatic with different levels of severity (mild, moderate and severe). Asymptomatic patients (n=5) were defined as positive for the SARS-CoV-2 PCR test, but no signs of diseases. The following criteria were used to stratify symptomatic SARS-CoV-2 patients in terms of the disease severity and accordingly with WHO guidelines (25); 1-MILD (n=18): individuals with no evidences of pneumonia; 2-MODERATE (n=6): individuals without need of invasive mechanical ventilation and without need of admission to hospital intensive care unit, but evidence of pneumonia and need of supplemental oxygen; 3-SEVERE (n=1): individuals with need of invasive mechanical ventilation and with need of admission to hospital intensive care unit. Patients demographic and relevant clinical data are summarized in table 1.

Healthy controls (n=5) are represented by volunteer individuals with no history of infection disorders that underwent CHUP for routine analysis.

All participants gave informed consent about all clinical procedures and research protocols were approved by the ethics committee of CHUP and CHVNG, Portugal.

### Human PBMCs isolation

Human peripheral blood mononuclear cells (PBMCs) from COVID-19 patients and healthy donors were isolated by gradient centrifugation using 1 volume of Lymphoprep™ (Stemcell Technologies) for 2 volumes of blood, 30 minutes at 900 x *g* with the brake off. The upper phase, containing the serum, was collected and stored at - 80°C. PBMCs (interphase) were collected, washed twice with PBS and incubated with FVD–APC-eFluor780 (eBioscience) for 30 minutes. Cells were washed with PBS, fixed with 2% formaldehyde (PanReac ApplieChem) and resuspended in FACS buffer (PBS 2%FBS). All procedures were performed under biosafety level 3 (BSL3) conditions.

### Flow Cytometry Staining

For lectin staining, cells were incubated with conjugated lectins (Vector Laboratories): Phaseolus Vulgaris leucoagglutinin (L-PHA-fluorescein, FITC), Galanthus Nivalis lectin (GNA-fluorescein, FITC), biotinylated Sambucus Nigra lectin (SNA) and biotinylated Aleuria aurantia lectin (AAL) for 15 minutes. Biotinylated lectins were incubated with streptavidin-PE-Cy7 (eBioscience) for 30 minutes. For surface marker staining, cells were stained for 30 minutes on ice while protected from light with the following antibodies: APC anti-human TCRγδ (clone B1), BV510 anti-human CD69 (clone FN50), PE anti-human CD25 (clone BC96), BV421 anti-human CD14 (clone 63D3), PE anti-human CD56 (clone HCD56), PE-Cy7 anti-human CD206 (clone 15-2) from Biolegend; eFluor™ 450 anti-human CD4 (clone RPA-T4), PerCP-eFluor™ 710 anti-human PD-1 (clone J105), PE-Cy5 anti-human CD19 (clone HIB19), eFluor™ 450 anti-human IgM (clone SA-DA4), eFluor™ 660 anti-human galectin 3 (clone M3/38), APC anti-human CD11c (clone BU15), PE-Cy5 anti-human CD86 (clone IT2.2) from eBioscience; PE anti-human TCRα/β (clone BW242/412) from Miltenyi; BV510 anti-human CD3 (clone OKT3) from BD Biosciences; for DC-SIGN staining, cells were incubated with DC-SIGN rabbit IgG (Biorad) followed by incubation with polyclonal swine anti-rabbit IgG (FITC; Dako) for 30 minutes on ice. Cells were resuspended in FACS buffer prior analysis. Data were obtained on a BD FACS Canto II instrument (Becton Dickinson) and analyzed using FlowJo v10.0 (Tree Star Inc.).

### *In vitro* assessment of glycan modulation of T cells

PBMCs from healthy and independent from the COVID-19 cohirt human donors were isolated from fresh collected blood, as described above. PBMCs were cultured in RPMI-1640 (Gibco) supplemented with 10% (v/v) heat-inactivated fetal bovine serum, 1% penicillin/streptomycin and 10% of plasma from 4 asymptomatic or moderate patients, for each condition. Different cell culture conditions were tested, namely human plasma concentration (25% and 10%) as well as time of culture (4 days, 2 days, 24h and 12h). The optimal culture condition (with lower cell death and stronger glycan modulation) was selected (10% human plasma and 24h cell culture). After 24h cell glycoprofile was evaluated using the same antibodies and lectin panel selected above. Data were obtained on a BD FACS Canto II instrument (Becton Dickinson) and analyzed using the FlowJo 10.0 software (Tree Star Inc.).

### Data visualization and statistical analysis

Data visualization and statistical analyses (non-parametric Mann-Whitney t-test) were done using GraphPad Prism 9 software.

The prediction capacity of DC-SIGN levels to discriminate patients who develop a poor disease course from those that have a good disease course was determined by plotting the receiver operating characteristic (ROC) curves and calculating the area under the curve (AUC). The cut-off that revealed the best balance between sensitivity and specificity was selected for the subsequent statistical analysis. Sensitivity, specificity, and positive and negative predictive values were calculated. Univariate binary logistic regression analysis was performed to test DC-SIGN levels and disease progression (good: asymptomatic maintained asymptomatic and/or severity decreased at least one category; poor: symptoms were not improved and/or escalate to a higher severity). In logistic regression, model goodness-of-fit was assessed by the Hosmer-Lemeshow statistic and test. Results are presented as odds ratios (ORs) for each category as compared with a predefined reference category, and their respective 95% confidence intervals (CIs). Odds ratios above one and below one are indicative, respectively, of higher and lower odds of develop poor disease course as compared with a reference category. Statistical analysis was performed using the statistical software SPSS version 25 (IBM Corp., IBM SPSS Statistics for Windows, Version 25.0, Armonk, NY; released 2017) The threshold used for statistical significance was *p*-value< 0.05.

## Results

### SARS-CoV-2 infected individuals display decreased β1,6-GlcNAc branched and α2,6-sialic acid *N*-glycans on peripheral T cells at diagnosis

Taking into consideration the fact that it is still unclear the underlying mechanisms that explain a differential inter-individual clinical presentation of COVID-19 at diagnosis, we herein characterized the glycosylation profile of T cells from patients’ peripheral blood. A lectin-based flow cytometry of the T cell populations was performed to evaluate glycosylation profiles. We started by analysing the levels of expression of ß1,6-GlcNAc branched *N*-glycan structures, known to have a major impact on the regulation of T cell activity and function(13, 16, 26), using the L-PHA lectin. Our results demonstrated that, overall, T cells from infected (IF) subjects have a decrease in branched *N*-glycan structures, particularly on CD8^+^ and γδ T cells, when compared to non-infected (non-IF) ones (Figure 1A, top and bottom left panel). Moreover, T cells from asymptomatic patients have lower levels of ß1,6-GlcNAc branched *N*-glycans, that gradually tend to increase along disease severity (**Figure 1A, right**). Notably, CD8^+^ T cells display a significant decrease in the levels of expression of complex branched *N*-glycans in asymptomatic and mild disease patients, when compared to non-IF individuals (**Figure 1A, top**).

**Figure 1.**
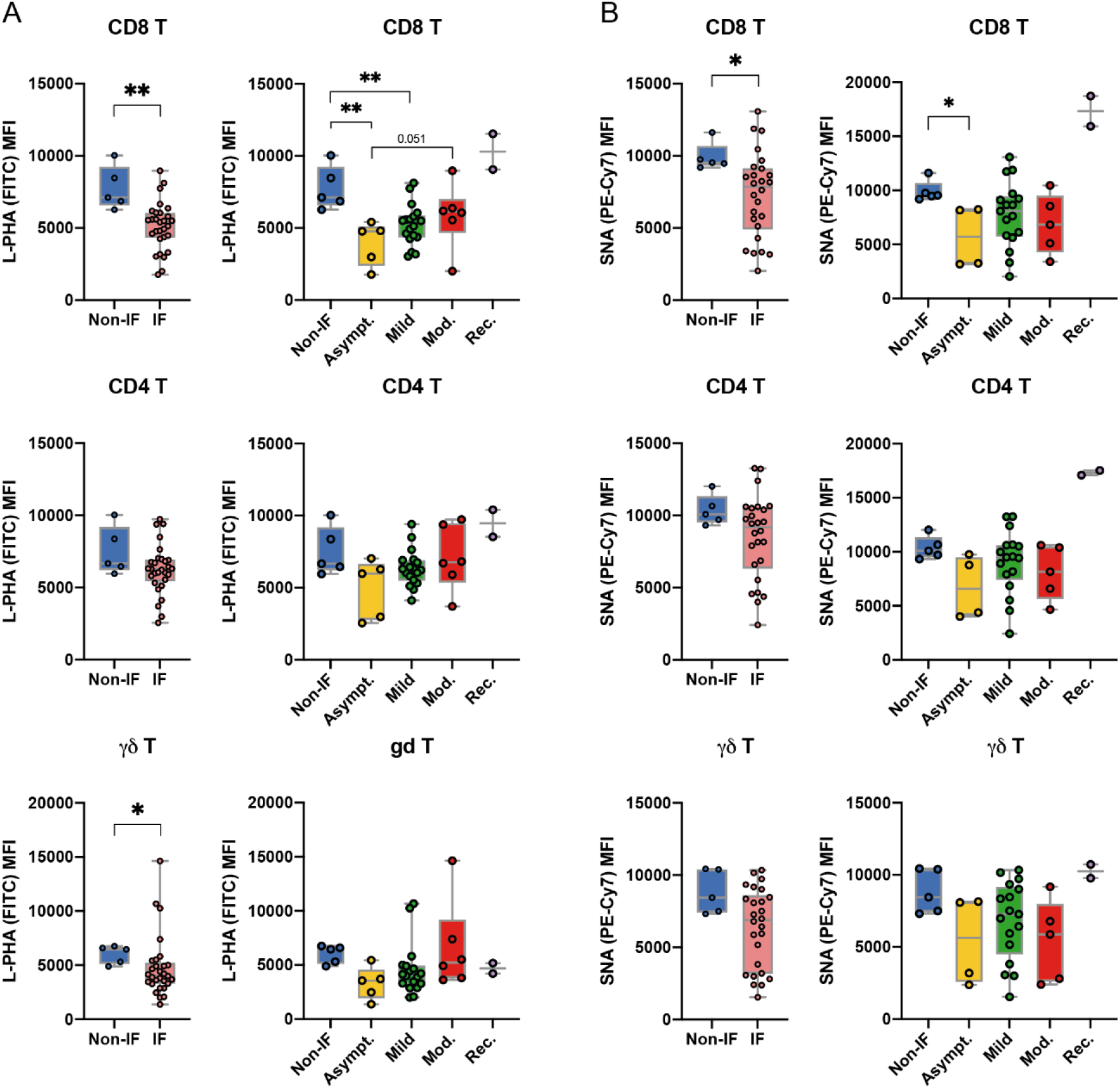
Peripheral T cell glycoprofile is altered upon SARS-CoV-2 infection at diagnosis. Levels (Mean intensity fluorescence, MFI) of (A) L-PHA lectin binding, detecting β1,6-GlcNAc branched *N*-glycans and (B) SNA lectin binding, detecting a-2,6 sialylation, in CD8^+^, CD4^+^ and γδ T cell subsets of non-infected donors (Non-IF; n=5) and infected (IF; n=30) patients. Levels of (A) L-PHA and (B) SNA lectin binding for each disease severity group: asymptomatic (Asympt.;n=5), mild (n=18), moderate (Mod.; n=6) and recovered (Rec; n=2) Each dot represents one patient. Mann-Whitney t-test was performed to evaluate statistically significance differences between each group-pair. * *p*-value<0.05, **<0.005.

Concerning α2,6-sialylation (recognized by the SNA lectin), already described to play a role in the regulation of T cells immune response(27), our results demonstrate that CD8^+^ T cells from IF patients display a significant decreased SNA binding, in comparison with non-IF individuals (**Figure 1B, top left**). Consistently, asymptomatic patients present the lowest levels of SNA binding compared to non-IF (Fig.1B, top right). Even though the levels of SNA binding were shown to be associated with age (**Supplemental Figure 1E**), there are no significant differences between the mean age of asymptomatic and symptomatic patients (**Supplemental Table 1**). Curiously, for both glycans, the two COVID-19 convalescent patients seem to recover (or surpass) the levels of expression of β1,6-GlcNAc branched and α2,6-sialic acid glycan structures in all T cell subsets (**Fig. 1A and B, right**). These results showed that CD8^+^T cells from asymptomatic patients display a defect in branched and sialylated *N*-glycans when compared with non-IF.

The differences observed in the glycosylation profile of T cells across severity, were not related to the abundance or activation of T cell populations, since CD3^+^, CD4^+^, CD8^+^ and γδ T cells showed similar frequencies as well as similar levels of PD1 and CD69 expression between the groups, with the exception of lowered CD3^+^ population and increased CD69^+^ within CD8^+^ T cells, in the mild disease group (**Supplemental Figure 1A, B and C**).

In addition, and since glycosylation is a dynamic process that can change along with disease status, we took advantage of a limited longitudinal follow-up analysis using a single patient, representative of each severity group (asymptomatic, mild, moderate and severe), which revealed that levels of expression of ß1,6-GlcNAc and α2,6-sialylation are restored in patients that improved disease severity at day 14 post-diagnosis **(Supplemental Figure 1D)**.

No major differences were observed for other glycan structures such as fucosylation (AAL binding) or mannosylation (GNA binding) in the different T cell subsets at diagnosis (**Supplemental Figure 1A and B**).

These results showed that SARS-CoV-2 infected individuals led to an *en bloc* decreased expression of specific glycan structures such as ß1,6-GlcNAc branched *N*-glycans and α2,6-sialic acid in T cells, predominately in CD8^+^ T cells and asymptomatic patients. The absence of differences on high mannose structures (GNA binding) or fucose residues on T cells (AAL binding), support the specificity of the glycan switch. In fact, decreased levels of β1,6-branched *N*-glycans in T cells have been pointed out as a mechanism of TCR threshold regulation, by hampering the TCR complex clustering and lowering the necessary amount of antigen-recognition to signal a cellular response(28). Moreover, surface α2,6-sialylation also regulates T cell immune response through galectins binding modulation(29–31).

In order to further clarify the underlying mechanism that imposes the changes in the glycosylation profile of circulating T cells upon SARS-CoV-2 infection, we have performed an *in vitro* assay, in which PBMCs isolated from healthy individuals were co-cultured with plasma from 4 patients from either each group: non-IF, asymptomatic or moderate disease. Our results showed that T cells, upon co-culture with plasma from asymptomatic patients displayed a significant decreased expression of β1,6-GlcNAc branched *N*-glycans (**Figure 2A**) and α2,6-sialylation (**Figure 2B**), when compared to T cells cultured with plasma from non-IF individuals and moderate disease patients. These results suggest the existence a specific serum factor that in asymptomatic patients drives a remarkable drop of complex branched and sialylated glycans structures on T cells. In fact, it is known that asymptomatic patients display a differential serum composition of cytokines and chemokines, when compared to symptomatic (32, 33). Moreover, T cell glycosylation is known to be modulated by specific interleukins, namely IL-2 and IL-7 that regulate the transcription of specific Golgi glycosyltransferases(17, 25), which renders the hypothesis that an extracellular factor could be mediating the T cells glycan switch in a more efficient manner in asymptomatic than symptomatic patients.

**Figure 2.**
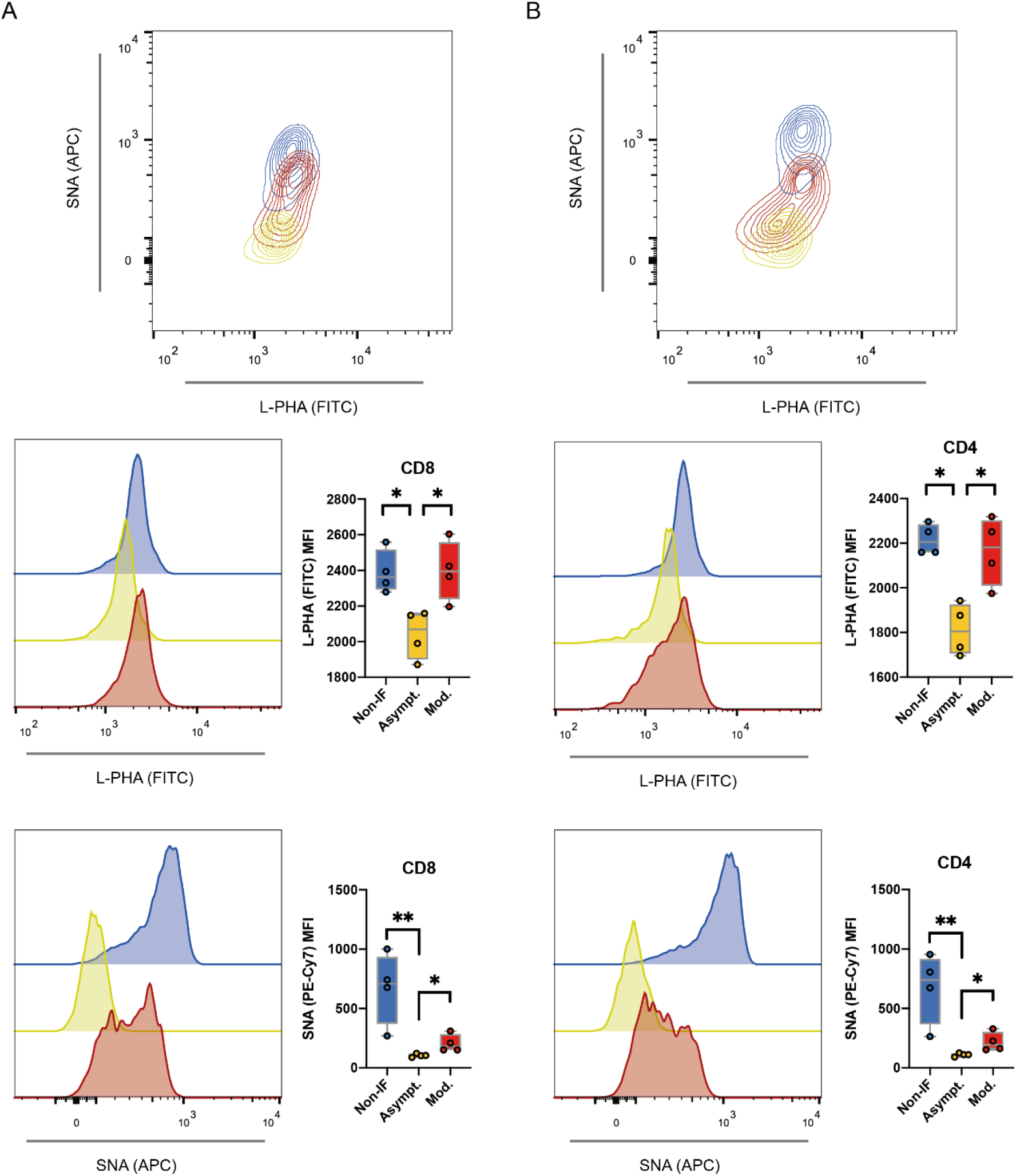
Patient-derived serum induces a differential T cell glycosylation comparing asymptomatic and symptomatic patients **(A)** L-PHA lectin and **(B)** SNA lectin binding (MFI) of CD8^+^ and CD4^+^ T cells from a healthy donor, independent of our cohort, incubated for 24-hours with 10% plasma from 4 patients from each group (Non-IF, asymptomatic or moderate). Each dot represents a subject. Gating strategies for each population are included in **Supplemental Figure 1A**. Mann-Whitney t-test was performed to evaluate statistically significance differences between each group-pair. * *p*-value<0.05, **<0.005.

### Levels of DC-SIGN expression in circulating monocytes predict COVID-19 prognosis

To gain further insights in the viral-glycan recognition mechanism, we have characterized the expression of specific GBPs expressed by innate immune cells that are known to sense and recognize specific viral glycans, instructing an immune response(34). Regarding the abundance of innate cells, we have observed an increase in the frequency of dendritic cells (DCs) as well as monocytes (Mo) in asymptomatic patients, when compared to Non-IF individuals, and no differences in terms of NK and NK-T cells subsets (**Supplemental Figure 1B**). Regarding their activation state, analysed by CD86 expression, only monocytes are shown to be activated in asymptomatic and mild disease patients (**Supplemental Figure 1C**). This is in accordance with previous findings(35).

Specifically, DC-SIGN, a C-type-lectin that recognizes mannose structures, is already known to play a role in the SARS-COV-2 recognition(24), being expressed by monocytes, immature DCs and macrophages. Our results showed that patients from the asymptomatic group displayed higher levels of DC-SIGN expression in monocytes compared moderate disease (**Figure 3A**), suggesting an increased capacity of asymptomatic patients to sense and recognize the virus.

**Figure 3.**
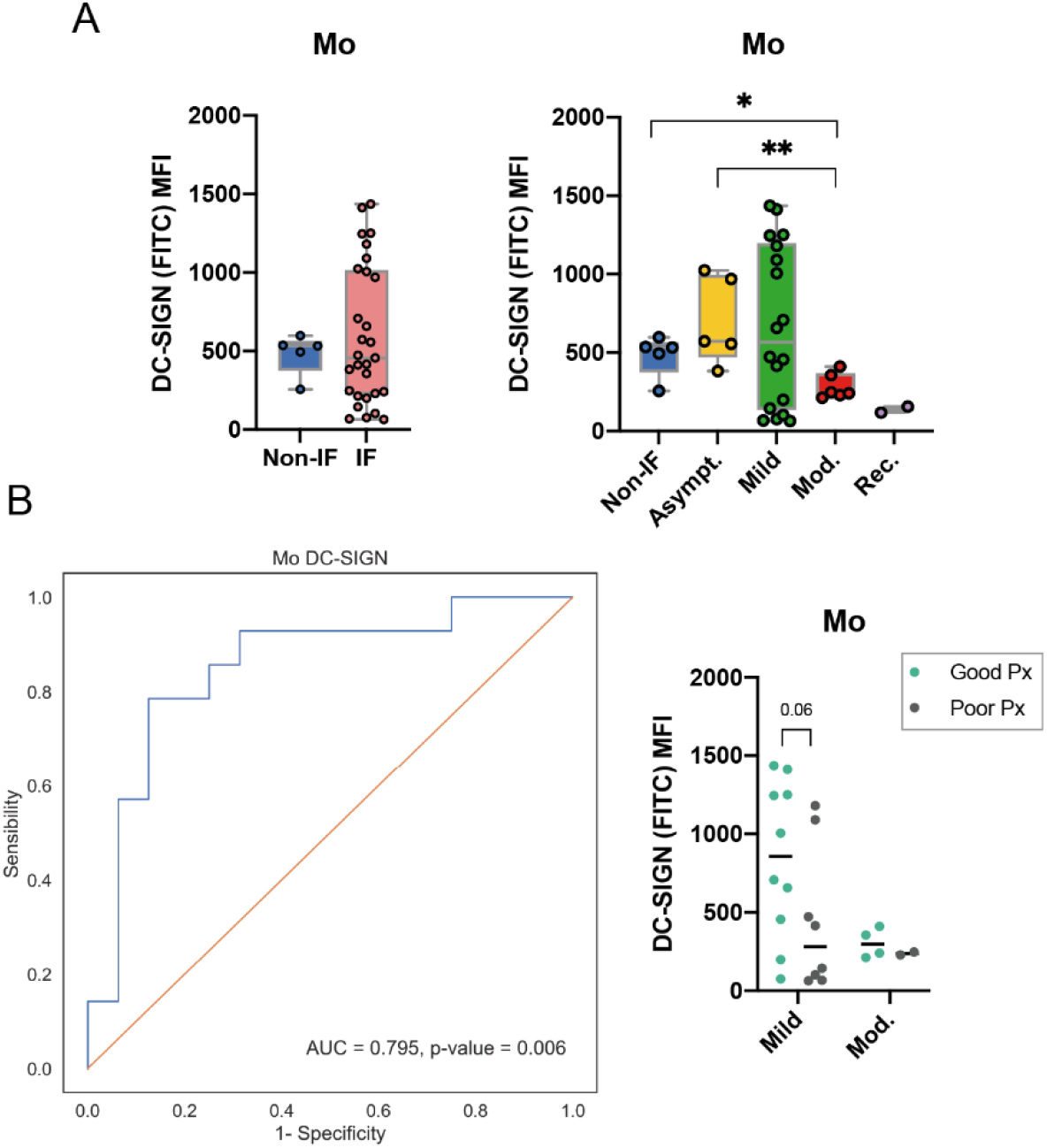
DC-SIGN expression in peripheral monocytes as a predictor of good vs. poor prognosis in COVID-19 patients. **(A)** Levels of DC-SIGN surface expression (MFI) in monocytes (Mo). **(B)** Receiver operating characteristic (ROC) curve plotted for the DC-SIGN expression levels in monocytes from COVID-19 patients. The distribution of DC-SIGN levels are represented regarding the prognosis, Px, (day 14; good Px in green and poor Px in grey) as well as the severity at diagnosis. Each dot represents one patient N=24. Gating strategies for each population are included in **Supplemental Figure 1A**. Mann-Whitney t-test was performed to evaluate statistically significance differences. * *p*-value < 0.05, ** < 0.005, or represented.

Taking into consideration the biological relevance of DC-SIGN expression in monocytes as the first sensing mechanism upon SARS-CoV-2 infection, and given the heterogenous expression of DC-SIGN among infected individuals (at diagnosis), (Figure 3A) we next analysed its prognostic value. A longitudinal cohort was used with clinical information in terms of disease progression (development of a poor *versus* good prognosis at day 14 post-diagnosis, characterized in Supplementary Table 1). Remarkably, the expression levels of DC-SIGN in monocytes were found to be able to stratify patients at diagnosis in terms of their likelihood of developing a good versus a poor disease prognosis with a specificity and sensitivity of 78.9% and 63.6%, respectively (**Figure 3B**). The univariate analysis demonstrated that high levels of DC-SIGN expression in monocytes at diagnosis increase the odds ratio (OD) associated with the prediction of having a good prognosis, OD = 6.548 *p*-value = 0.011. In fact, the recognition of the coronaviruses’ glycans by innate immune cells was demonstrated to be mediated by DC-SIGN(23, 24). An increased expression of this C-type-lectin at the initial days of infection suggests a more efficient viral recognition and subsequent clearance, resulting in a good prognosis of the disease.

## Discussion

Our results demonstrated for the first time that SARS-CoV-2 infection imposes a glycosylation reprogramming of adaptive immune cells suggesting a specific glycan switch of T cells that may define their ability to successfully deal with infection. We identified a specific glycosylation signature of circulating T cells that is associated with their activity and function, distinguishing infected patients from non-infected individuals. Our results also suggest that the glycan switch imprinted in circulating T cells could be mediated by a serum extracellular factor(s), occurring just after SARS-CoV-2 infection (**Supplemental Figure 2**). In fact, previous evidences from others and us demonstrated that the deficiency in branched *N*-glycan structures on T cells imposes a hyper-reactive phenotype with decreased threshold of T cell activation and increased T cell activity(13, 28). We herein propose that the immune response against SARS-CoV-2 infection appears to be influenced by the glycosylation profile of circulating T cells which defines their effective functions. In fact, a pronounced deficiency of complex branched and sialylated glycans on CD8^+^ and γδ T cells were observed in asymptomatic COVID-19 patients. This dynamic and plastic glycoimmune modulation (“Glyco-Immune Alert” mechanism) may constitute a novel mechanism of host-response, that contributes to further understand the immunological differences among infected individuals(36).

Furthermore, we also showed another glycan-based mechanism of host response to SARS-CoV-2 driven by the upregulation of the viral’s glycans recognizing receptor DC-SIGN in monocytes, with prognostic application when detected at diagnosis.

This study unlocks the identification of a specific glycosignature of T cells as well as a prognostic biomarker in COVID-19. Further studies are needed to explore the mechanistic impact of the T cell glycan switch in infection and along disease course. Moreover, it is fundamental to identify the critical serological factor(s) that instructs this T cells glyco-reprogramming, as a potential new biomarker and therapeutic target. Our study was conducted during the first wave of COVID-19 (March-July 2020) having a limitation in terms of sample size, indicating the need to validate these promising observations in larger and well-characterized multicentric cohorts as well as analysing the impact of other SARS-CoV-2 variants in T cells glycan switch.

These new evidences in COVID-19 pave the way to the identification of a specific blood glycosignature able to stratify patients at diagnosis according with their risk to evolve to worsen disease. This will certainly contribute to improve vaccination strategy and patients risk stratification, optimizing an effective allocation and management of health care resources such as ventilators and intensive care facilities.

## Data Availability

Data is not available.

## Acknowledgments

We would like also to thank to all the Portuguese clinicians involved in COVID-19 patients ‘care, with a special thanks to those from CHUP and CHVNG that directly or indirectly contributed to this work (Dr. Tiago Teixeira from CHVNG) and to all the patients that accepted to participate in this study. We also acknowledge the nurses and technicians that collaborated in the collection of the samples, especially Nurse Teresa Cruz from CHUP.

## Author contributions

IA, MV and SSP designed research. IA, MV, JG, AF, AMD and CSR performed research. SSP contributed new reagents/analytical tools. NS, LM, MAA and RSC provided and characterized clinical samples. IA and MV analysed data. IA, MV and SSP wrote the manuscript with contributions from all authors.

