## Supplemental Fig. 1 and Table 1 for "SARS-CoV-2 infection drives a glycan switch of peripheral T cells at diagnosis"

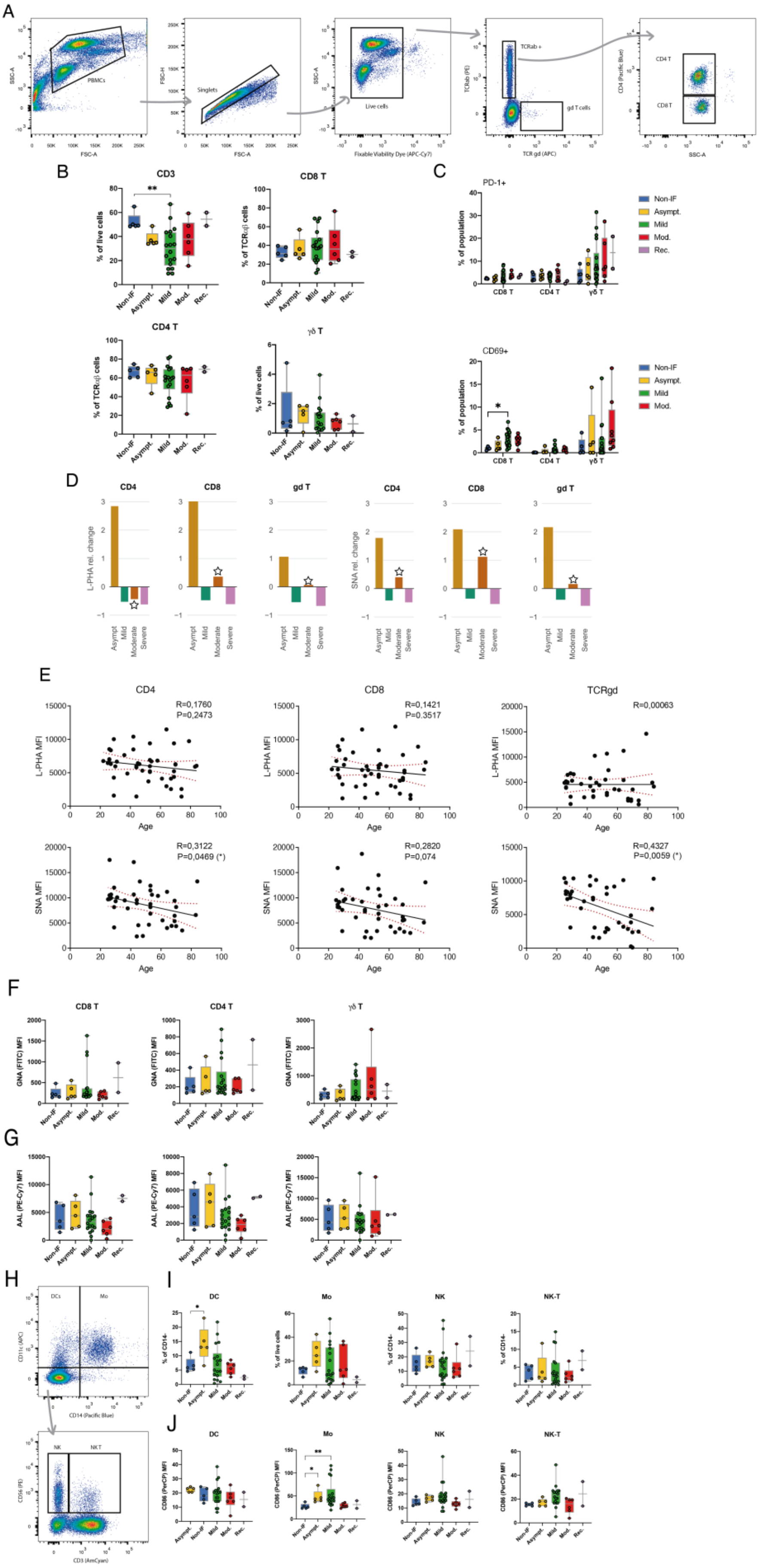

A
PBMCs
Singlets
Live cells
TCRab +
gd T cells
CD4 T
CD8 T
FSC-A
SSC-A
FSC-H
Fixable Viability Dye (APC-Cy7)
TCR gd (APC)
B
CD3
CD8 T
CD4 T
γδ T
% of live cells
% of TCRαβ cells
Non-IF
Asympt.
Mild
Mod.
Rec.
C
PD-1+
CD69+
% of population
D
CD4
CD8
gd T
L-PHA rel. change
SNA rel. change
Asympt
Mild
Moderate
Severe
E
CD4
CD8
TCRgd
R=0,1760
P=0,2473
R=0,1421
P=0.3517
R=0,00063
R=0,3122
P=0,0469 (*)
R=0,2820
P=0,074
R=0,4327
P=0,0059 (*)
L-PHA MFI
SNA MFI
Age
F
GNA (FITC) MFI
G
AAL (PE-Cy7) MFI
H
DCs
Mo
NK
NKT
CD14 (Pacific Blue)
CD3 (AmCyan)
I
DC
Mo
NK
NK-T
% of CD14-
% of live cells
J
CD86 (PerCP) MFI

**Supplemental Figure 1 – (A)** Gating strategy for multicolour flow cytometry analysis; **(B)** Frequency of $CD3^+$, $CD8^+$, $CD4^+$ and γδT cells in peripheral blood from non-IF individuals and patients grouped according severity of disease. **(C)** PD1- and CD69-expressing T cell frequencies of peripheral CD4, CD8 and γδT cells from non-infected and healthy donors (Non-IF), and COVID-19 patients, at the time of diagnosis. **(D)** Relative change of L-PHA and SNA binding levels in several T cell subtypes at day 14 post-diagnosis, compared to the analysis at diagnosis, in N=1 patient/group (asymptomatic; mild and moderate); ☆ in the moderate group (orange bar) highlights a patient that has improved, with a recovery in disease severity at day 14 post-diagnosis, from moderate to mild disease, which was accompanied with a delayed T cell glycan switch. Patient with mild disease (green bar) maintained the disease status 14 days after diagnosis. **(E)** Correlation of the age from the cohort-included subjects and levels of L-PHA or SNA in the different T cell subsets analysed, with respective P-value as well as correlation coefficient, R. **(F)** Levels of GNA binding (MFI) and **(G)** AAL binding in the non-infected donors (Non-IF), asymptomatic (Asympt), mild and moderate patients in $CD8^+$, $CD4^+$ and γδT cells. **(H)** Gating strategy for multicolour flow cytometry analysis. **(I)** Frequency and activation of dendritic cells (DC), monocytes (Mo), natural killer cells (NK) and natural killer T cells (NK-T) of non-IF and COVID-19 patients at time of diagnosis. **(J)** Cellular activation measured by the expression levels of CD86 of respective innate immune cells of non-infected subjects (non-IF) and asymptomatic, mild, moderate and recovered COVID-19 patients. Each dot represents one patient. Mann-Whitney test was performed to evaluate statistically significance differences. * $p$-value < 0.05, ** < 0.005.

**Supplementary Table 1**: Characterization of COVID-19 cohort in terms of clinical and epidemiological parameters. Data on age, gender and existence of comorbidities were obtained at diagnosis. Clustering of patients either by Good (asymptomatic who remained with no symptoms and amelioration of symptomatology, at day 14) or Poor (worsen severity at day 14) disease course.

| Characteristics | Severity COVID-19 | | | Disease Course | |
|---|---|---|---|---|---|
| | Asymptomatic (N=5) | Mild (N=19) | Moderate (N=6) | Good (N=19) | Poor (N=13) |
| **Age (y)** | 57,4 ± 14,32 | 51,24 ± 14,18 | 63,17 ± 11,89 | 51,11±11,67 | 55,50±17,50 |
| **Age groups (y)** | | | | | |
| 22-65 | 3(60%) | 16(84,21%) | 4(66,67%) | 15(78,95%) | 9(69,23%) |
| >65 | 2(40%) | 3(15,79%) | 2(33,33%) | 4(21,05%) | 4(30,77%) |
| **Gender** | | | | | |
| Male | 1(20%) | 8(42,11%) | 4(66,67%) | 8(42,11%) | 4(30,77%) |
| Female | 4(80%) | 11(57,89%) | 2(33,33%) | 11(57,89%) | 9(69,23%) |
| **Coexisting comorbidities** | | | | | |
| Yes | 1(20%) | 6(31,58%) | 5(83,33%) | 4(21,05%) | 10(76,92%) |
| No | 4(80%) | 13(68,42%) | 1(16,67%) | 15(78,95%) | 3(23,08%) |
| **Disease Progression** | | | | | |
| Good (improved/asymp) | 5(100%) | 10(52,63%) | 3(50%) | -- | -- |
| Bad (worsen severity) | 0 | 9(47,37%) | 3(50%) | -- | -- |
