## Supplemental Fig. 2 for "SARS-CoV-2 infection drives a glycan switch of peripheral T cells at diagnosis"

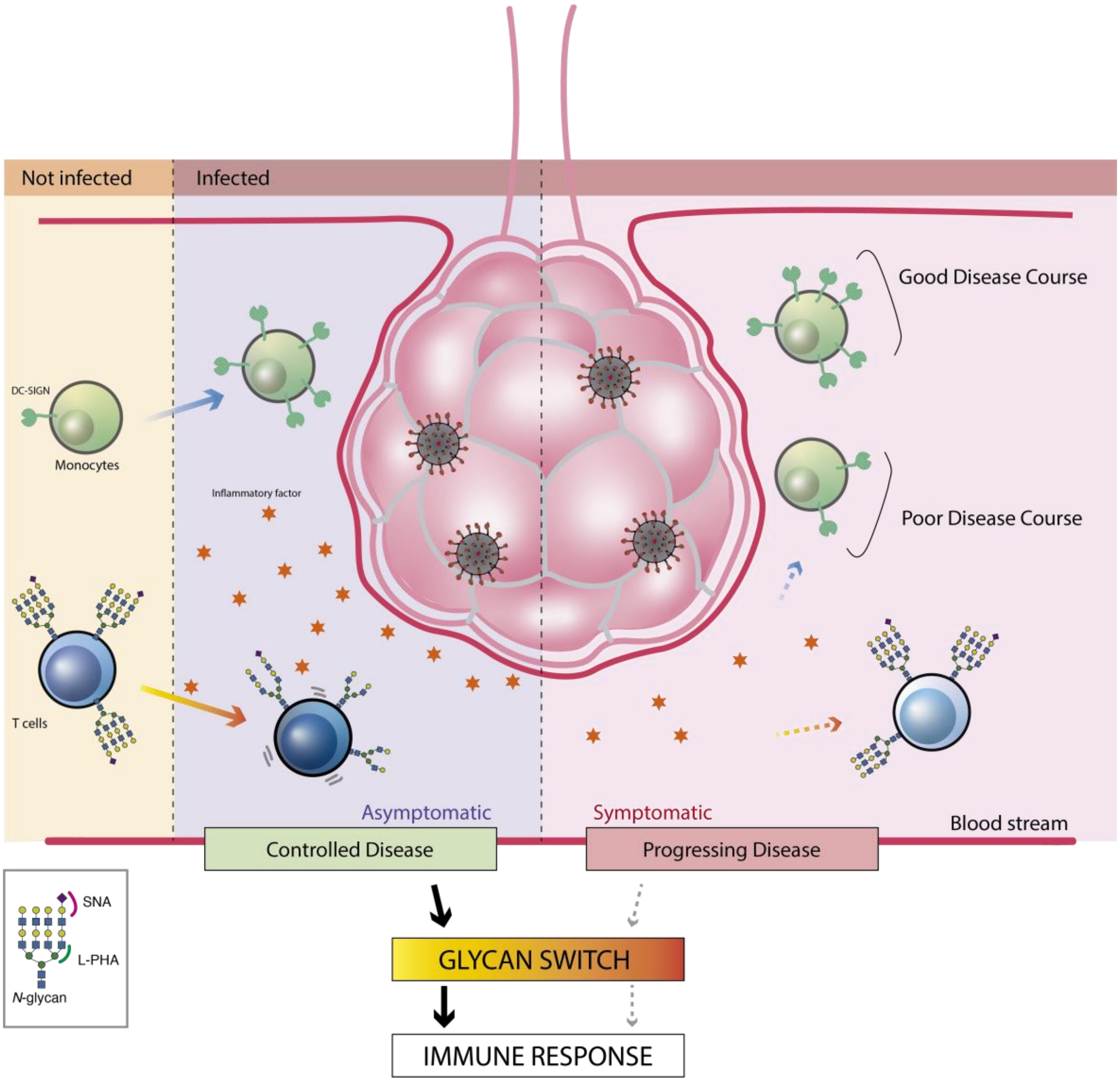


**Supplemental Figure 2** - SARS-CoV-2 infection results in changes in circulating T cells glycoprofile together with alterations in monocytic DC-SIGN expression that are correlated with COVID19 symptomatology and disease course. Non-infected (Non-IF) individuals display high levels of ß1,6-GlcNAc branched *N*-glycans and sialylation on T cells surface, and low levels of DC-SIGN expression in monocytes, which is related with immune homeostasis. Upon infection, two main types of responses occur in individuals: development of symptoms (symptomatics) or not (asymptomatics). Asymptomatic status was correlated with the existence of a glycan switch of peripheral T cells, characterized by a specific glycosignature on T cell surface not found in symptomatic patients. The decrease on complex branched and sialylated N-glycans on T cells appear contribute to the T cell priming and activation by lowering thresholds of TCR signalling in asymptomatics. Moreover, we found an upregulation of DC-SIGN in monocytes, which has been related to SARS-CoV-2 glycans' recognition, a feature that reflects an efficient virus detection system. Patients that develop symptoms and a progressing disease did not display a significant glycosylation modulation of circulating T cells, which we propose to be related to an inefficient immune response towards SARS-CoV-2. These patients also exhibit a failure in the upregulation of DC-SIGN in monocytes that was demonstrated to be associated with disease prognosis.
